# Epidemiology of intestinal helminthiasis with an emphasis on taeniasis in the Chipata district of the Eastern province of Zambia

**DOI:** 10.1101/2023.08.01.23293479

**Authors:** Gideon Zulu, Chummy Sikasunge, Tamara M. Welte, Martin Simunza, Dominik Stelzle, Veronika Schmidt, Alex Hachangu, Wilbroad Mutale, Maxwell Masuku, Mwelwa Chembensofu, Clarissa Prazeres da Costa, Kabemba E. Mwape, Andrea S. Winkler, Isaac K. Phiri

**Author notes:** Corresponding author. (GZ). These authors contributed equally to this work.

## Abstract

**Background:** Intestinal helminth infections are among the most common infections worldwide and have a negative impact on the health, education, nutrition and economic development of affected populations. This study aimed to estimate the prevalence of intestinal helminthiasis, including *T. solium* taeniasis, using a large-scale community-based study in the Chiparamba area of Chipata District of the Eastern province of Zambia.

**Methods/Principle Findings:** A cross-sectional study was conducted between June 2019 and December 2022 in a rural community of 25 randomly selected villages known to be at risk for *T. solium* infection. Stool samples were examined for intestinal helminths using the formol-ether concentration technique and further tested for taeniasis by copro antigen-ELISA (copro Ag-ELISA). Descriptive statistical analyses were conducted, and associations between the disease prevalence of active infections and individual- and village-level variables were determined using the chi-square or Fisher’s exact test. Predictors of an individual being positive for either taeniasis or other soil-transmitted helminths were determined using binary logistic regression. A total of 2762 stool samples were examined. One hundred ninety-five (7.1%) tested positive for at least one helminthic parasite on microscopy, with hookworm being the most frequent (84, 3.0%), followed by *S. mansoni* (66, 2.4%). For taeniasis, 11 (0.4%) participants were positive for *Taenia* spp. microscopically, while 241 (8.7%) tested positive via copro Ag-ELISA. On bivariate analysis, male sex was significantly associated with the prevalence of intestinal parasites (*p* = 0.012) but not with that of taeniasis based on copro Ag-ELISA results. Village level differences were significant for infection with intestinal helminths as well as for taeniasis positivity on copro Ag-ELISA (*p* <0.001).

**Conclusion:** Intestinal helminths, including *T. solium* taeniasis, are prevalent in the Chiparamba area of the Chipata district in the eastern province of Zambia, supporting the clear need for further targeted public health interventions for surveillance and control.

**Author summary:** Intestinal helminth infections including schistosomiasis and *T. solium* are among the major neglected tropical diseases affecting communities with poor access to clean water, sanitation and hygiene. *T. solium* taeniasis is also associated with communities practicing free range pig management. These infections are common in Sub-Saharan Africa, South America, and Asia, where they negatively impact on the health, education, nutrition and economic development of the affected population. Our study aimed to estimate the prevalence of intestinal helminthiasis, including *T. solium* taeniasis in the Chiparamba area of Chipata district of the Eastern province of Zambia. We found that 7.1% of the stool samples examined microscopically, were infected with at least one intestinal parasite. The most common parasites found were hookworm (3.0%) and *S. mansoni* (2.4%) while 0.4% were *Taenia* spp. We also found that 8.7% of the stool samples examined were positive for *Taenia* spp. antigens. Males were more associated with having intestinal parasites. Village level differences for infection with intestinal helminths as well as being positive for *Taenia* spp. antigens were also observed. The study shows that intestinal helminths including *T. solium* are present in our study community and require public health interventions for surveillance and control.

## Introduction

Intestinal helminth infections are among the most common infections worldwide, with an estimated 1.5 million people infected globally [1]. These infections are common in Sub-Saharan Africa (SSA), South America, China and Asia and affect communities with poor access to clean water, sanitation and hygiene [1]. The most frequently found soil-transmitted helminths (STH) include hookworms, *Trichuris trichiura, Ascaris lumbricoides*, *Strongyloides stercoralis* and *Schistosoma* spp. These infections are of public health importance because of their negative impact on the health, education, nutrition and economic development of the affected population [2,3]. Intestinal helminth infections may result in malnutrition, iron-deficiency anaemia, and malabsorption. In addition, stunted growth and delays in cognitive development are frequent consequences of heavy intestinal helminth infection [4]. STH and *Schistosoma* spp. have therefore been targeted for control and eradication [5].

Schistosomiasis affects approximately 240 million people worldwide, and the majority of these people are found in sub-Saharan Africa, with more than 200 000 deaths per year. Humans that come into contact with bodies of water containing fresh water snails are at the highest risk of infection [6].

*Taenia solium* is another intestinal helminth that is known to have serious public health and economic impacts in low-income and middle-income countries (LMICs), particularly in communities where sanitation is poor, pigs are reared through the free-range system, and meat inspection is not practiced [7,8]. Humans and pigs are both affected by *T. solium* [9], and both may develop cysticercosis when they ingest *T. solium* eggs. Pigs acquire *T. solium* eggs mainly through consumption of human faeces from a *T. solium* tapeworm carrier, while humans become infected either directly through the faecal oral route (autoinfection) or indirectly through consumption of contaminated food or water. Humans develop tapeworm infection (*T. solium* taeniasis) when they consume raw or undercooked pork.

Although *T. solium* taeniasis has been recognized as a serious and emerging challenge to public health in Africa [5], there are limited data on its incidence and prevalence in most endemic areas due to a lack of adequate surveillance, monitoring and reporting systems [10,11]. Within Zambia, circulating antigens (Ag-ELISA) for porcine cysticercosis (PCC) were detected in 15% to 34% of pigs in Eastern, Western and Southern provinces [8,12]. In the two districts Katete and Sinda of the Eastern province, prevalences of 46% and 68%, respectively, were detected in slaughter-aged pigs based on full carcass dissection [13]. Within the same province, human taeniasis prevalence based on copro Ag-ELISA was found to range from 6.3% to 12% [7,14]. Studies on intestinal helminths previously conducted in the Southern province of Zambia found prevalences of schistosomiasis and STH among adults of 13.9% and 12.1%, respectively [15]. Most studies on intestinal helminths in Zambia were conducted in children [16–18]. Within the Eastern province, no studies have been performed to estimate the prevalence of intestinal helminths, including schistosomiasis, and previous studies on human *T. solium* infections were conducted with a small number of participants or villages. This study aimed to estimate the prevalence of intestinal helminths with a focus on taeniasis in a large-scale community-based study in the Chiparamba area of Chipata District.

## Materials and methods

### Study site

The study was conducted in the Chiparamba area of the Chipata district of the Eastern province of Zambia. Chiparamba is located approximately 50 km from the central business district of Chipata (Fig 1). The study was conducted between June 2019 and December 2022.

**Fig 1:**
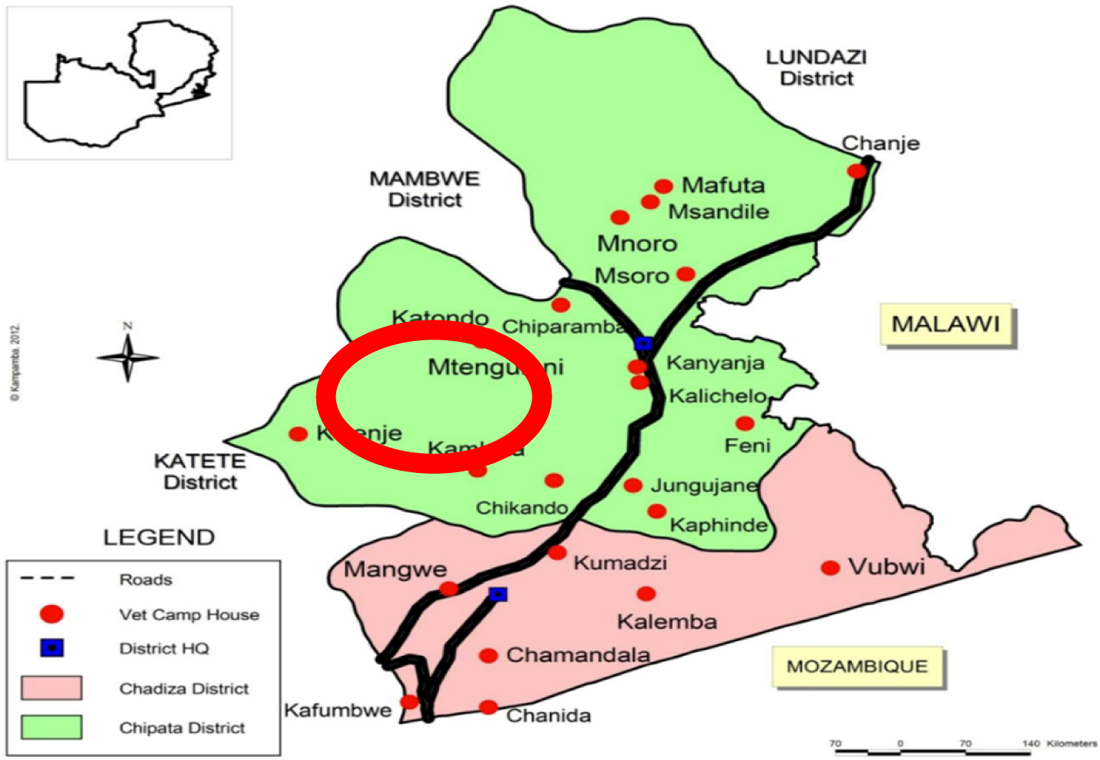
Map showing the Chiparamba area of Chipata district, Eastern province, Zambia. The red circle signifies the study area.

The people of Chiparamba mainly practice subsistence farming with cattle, goats, pigs and chickens as the main livestock kept. The main crops grown include maize, groundnuts, bananas and cotton. Hand pumps are the main source of water for most villages. Their homes are made of mud bricks, and few of them have latrines. Since sanitation is poor, pigs access human faeces in nearby bushes that are used as latrines by the villagers. Later, these pigs are sold on the local market by the villagers. For this study, Chipata district was chosen based on our findings of a high prevalence of taeniasis and cysticercosis in the neighboring district of Katete among people keeping free-range pigs [7,12,13].

### Study design and participant selection

This study was part of the TEPIM project “Establishment and application of a *Taenia solium* Experimental Pig Infection Model and investigation of environmental factors associated with transmission of *T. solium* in endemic villages of the Eastern and Southern provinces of Zambia”. Here, we report the human part of this study, which involved the collection of tapeworm proglottids to be used for the pig infection model. A cross-sectional study was conducted in the Chiparamba area of the Chipata district. A list of all the villages under the Chiparamba Rural Health Centre (RHC) catchment was obtained from the RHC, and villages were randomly selected for the study. Prior to sampling, the community was informed about the study and the activities that were to be conducted during a community sensitization day. To be eligible, study participants had to be willing and able to participate in all aspects of the study, including providing stool samples and informed consent (signature or thumbprint with impartial witness; assent for minors with parental consent), living in the study area and aged 10 years or older. Participants were excluded if the listed inclusion criteria were not met or if they were seriously ill (unable to engage in the normal activities of daily living without assistance because of their illnesses) at the time of the study.

### Sample size

Sample size estimation was based on the number of *T. solium* proglottids required for the experimental pig infection study. A total of 40 proglottids were required for the experimental infection study. At a reported taeniasis prevalence of 0.3% by microscopic examination [14] and assuming that two gravid proglottids were to be obtained from each tapeworm collected, a total of 6,668 people were to be tested for taeniasis from two districts, Chipata and Gwembe. Because the experimental infection study was discontinued due to logistical reasons and the Gwembe district site was not sampled, a sample size of 3,000 participants was targeted to estimate the prevalence of taeniasis and STH from the Chipata district.

### Sample collection and storage

Information on the age, sex and village of each recruited participant was collected. Recruited individuals were each given a stool sample bottle. They were asked to fill at least half of the sample bottle with specimen and submit the samples to the research teams amounting to up to 30 g of stool. As soon as the stool samples were submitted to the research team by the participants, they were placed on ice in a cooler box and stored in a refrigerator. Within 24 hours of collection, the stool samples were aliquoted. Each sample was placed in 10% formalin to allow later analyses for the diagnosis of taeniasis. The formalin aliquots were kept at room temperature until analysis.

### Stool sample examination by microscopy

Microscopic examination of stool was performed to determine the presence of *taenia* eggs and other intestinal helminths using the formol-ether concentration technique [19,20]. In brief, 2 g of fresh stool was transferred into a centrifuge tube containing 8 ml of 10% formal saline solution and thoroughly mixed using bamboo skewers. Two mills of ether were then added, and after closing the tubes with a stopper, vigorously shaken for thorough mixing. The tubes were then centrifuged at 2500 rpm for 5 min. The supernatant fluid was decanted, leaving the sediment in the tube. A drop of the sediment was obtained and placed on a slide. A cover slip was then placed, and the slide was examined under the microscope. Duplicate smears were made for each sample to enhance sensitivity. The slide was then scanned systematically using the 10X objective, and if anything suspicious was seen, the 40X objective was used for a more detailed examination. The presence of a *taenia* egg on a slide was recorded as being positive for taeniasis, and the presence of other parasite eggs was also recorded during the examination.

### Stool sample examination by copro-antigen enzyme-linked immunosorbent assay (Copro-Ag ELISA)

The stool samples were analysed for the presence of copro-antigens using a polyclonal antibody-based antigen-ELISA (copro Ag-ELISA) as described by Allan et al 1990 [21] and with modifications as suggested by Mwape et al 2012 [14]. In brief, an equal amount of stool sample and phosphate-buffered saline (PBS) were mixed. The mixture was allowed to soak for one hour with intermediate shaking and thereafter centrifuged at 2000 × g for 30 minutes. The supernatant was then used for the Ag-ELISA. Polystyrene ELISA plates (NuncH Maxisorp) were then coated with the capturing hyperimmune rabbit anti-Taenia IgG polyclonal antibody diluted at 2.5 mg/ml in carbonate-bicarbonate buffer (0.06 M, pH 9.6). After coating, the plates were incubated for 1 hour at 37°C and then washed once with PBS in 0.05% Tween 20 (PBS-T20). All wells were then blocked by adding blocking buffer (PBS-T20+ 2% New Born Calf Serum) and incubated at 37°C for 1 hour, after which 100 ml of the stool supernatant was added, and the plates were incubated for 1 hour at 37°C followed by washing with PBS-T20 five times. One hundred microliters of biotinylated hyperimmune rabbit IgG polyclonal antibody diluted at 2.5 mg/ml in blocking buffer was added as a detecting antibody, and the plates were incubated for 1 hour at 37°C followed by washing five times. One hundred microliters of streptavidin-horseradish peroxidase (Jackson ImmunoResearch Lab, Inc.) diluted at 1/10,000 in blocking buffer was added as a conjugate and incubated at 37°C for 1 hour. After washing five times, 100 ml of Ortho Phenylene Diamine (OPD) substrate, prepared by dissolving one tablet in 6 ml of distilled water and adding 2.5 ml of hydrogen peroxide, was added. The plates were incubated in the dark for 15 minutes at room temperature, after which 50 ml of sulfuric acid (4 N) was added to each well to stop the reaction. The plates were read using an automated spectrophotometer at 490 nm with a reference of 655 nm. To determine the test result, the optical density (OD) of each stool sample was compared with the mean of a series of 8 reference Taenia-negative stool samples plus 3 standard deviations (cut-off).

### Data processing and analysis

Data were entered electronically via tablets into the Kobo toolbox, an application for data collection installed on tablets [22]. The data were stored in a Microsoft Excel 2016 database (Microsoft Corporation; Redmond, WA, EUA), and later, statistical analyses were performed using IBM SPSS Statistics (Version 23). Descriptive analyses were conducted, and to determine associations between the categorical variables, a bivariate analysis was performed using either the chi-square or Fisher’s exact test. Predictors of an individual being positive for either taeniasis or STH were determined using binary logistic regression. The variables included in the model were age, sex, village and results of the copro antigen-ELISA and microscopy examination. A significant Omnibus Test of Model Coefficients (*p* ≤ 0.050) and a nonsignificant Hosmer‒Lemeshow test (*p* > 0.050) were used to determine if the model fit the data [23,24]. The model returned the odds ratios with their 95% confidence intervals for taeniasis and other intestinal helminth positivity. All statistics were considered significant at *p* ≤ 0.05.

### Ethical approval

Ethical clearance was obtained from the ERES CONVERGE Institutional Review Board (IRB) (Reference number 2018-March-002). Approval was also obtained from the Ministry of Health through the Zambia National Health Research Authority (ZNHRA). Further approval was sought from the community leaders before commencement of the study. All participants were informed about all aspects of the study before inclusion, and all signed an informed consent form. For individuals below the consenting age, written formal consent was obtained from their parents or guardians.

## Results

A total of 2,921 participants were recruited in the study, of which 2,762 (95%) submitted stool samples for analysis. Of those who submitted stool samples, the majority (1,610, 58.3%) were female (Fig 2). The minimum and maximum ages were 10 and 100 years, respectively, with a mean age of 29 years (SD = 17.5). The majority (2,074, 75%) of the participants were younger than 40 years, with the age group 10 – 20 years contributing the most (Table 1).

**Fig 2.**
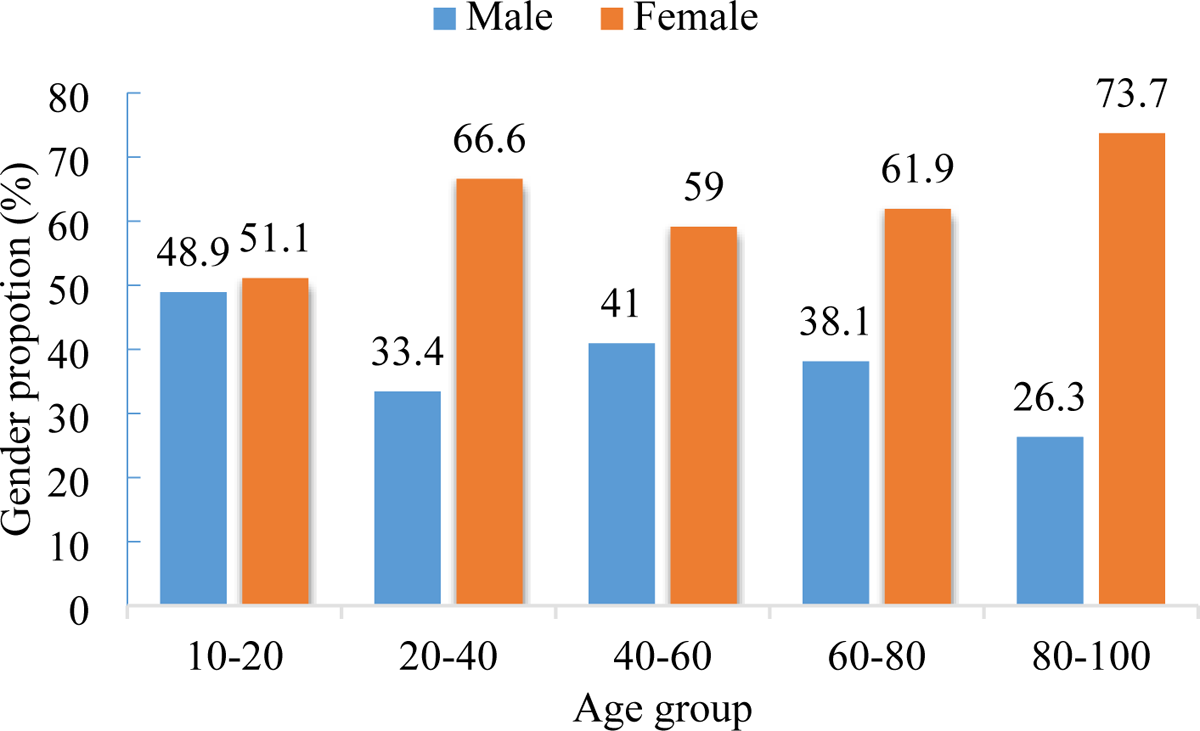
Sex distribution by age group of participants in the Chiparamba area of Chipata district, Eastern province, Zambia.

**Table 1.** Distribution of participants by age group in the Chiparamba area of Chipata district, Eastern province, Zambia.

|  |  | Number | Percent |
| --- | --- | --- | --- |
| Age group | 10-20 | 1199 | 43.4 |
|  | 20-40 | 875 | 31.7 |
|  | 40-60 | 514 | 18.6 |
|  | 60-80 | 155 | 5.6 |
|  | 80-100 | 19 | 0.7 |

### Intestinal helminths

One hundred ninety-five participants (7.1%, 95% CI = 6.2 – 8.1) tested positive for at least one intestinal parasite on microscopy. Twelve (0.4%, 95% CI =0.2 – 0.7) participants had more than one parasite. Overall, seven different species of intestinal parasites were identified. These included *Taenia* spp, *S. mansoni, A. lumbricoides*, *T. trichiura, Enterobius vermicularis, Entamoeba* spp. and hookworm spp. (Table 2). The most frequent infection was due to hookworm (84, 3.0%), followed by *S. mansoni* (66, 2.4%). Infections by *T. trichiura* (11, 0.4%), *A. lumbricoides* (7, 0.3%), *E. vermicularis* (4, 0.1%), and *Entamoeba* spp. (6, 0.2%) were less prevalent (Table 2). Intestinal helminth infection was higher in the 20 – 40 and 40 – 60-year-old age groups, although this difference within age categories was overall not statistically significant (*p* = 0.900) (Fig 3, Table 3). There was a statistically significant difference (*p* = 0.012) in the prevalence of infection between sexes, with more males infected (Table 3). Among villages, the prevalence of intestinal helminths was also significantly different (*p* < 0.001), with prevalence ranging from 1.8% in Mzamo village to 20.8% in Kochiwe village (Table 4).

**Fig 3.**
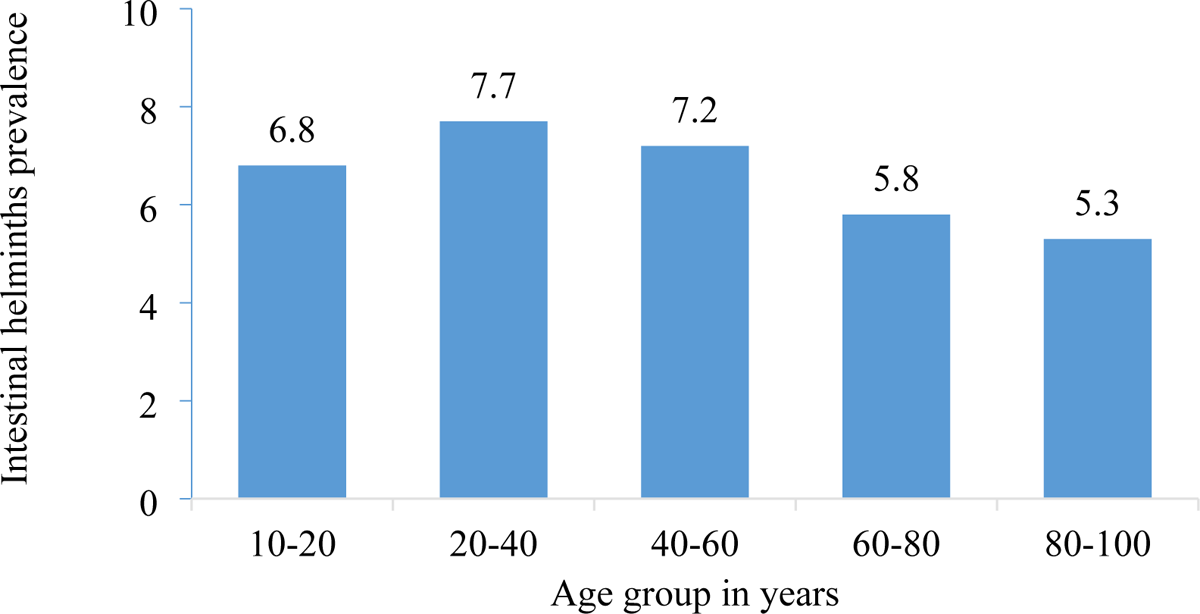
Prevalence of intestinal helminths by age group in the Chiparamba area of Chipata district, Eastern province, Zambia.

**Table 2.** Intestinal parasites identified through microscopy in the Chiparamba area of the Chipata district, Eastern province, Zambia. (n = 2762)

|  |  | Positive | Percent | 95%CI |
| --- | --- | --- | --- | --- |
| Single infections | *Taenia* spp. | 11 | 0.4 | 0.2 – 0.7 |
|  | Hookworm | 84 | 3.0 | 2.4 – 3.7 |
|  | *Ascaris lumbricoides* | 7 | 0.3 | 0.2 – 0.6 |
|  | *Trichuris trichiura* | 11 | 0.4 | 0.2 – 0.7 |
|  | *Enterobius vermicularis* | 4 | 0.1 | 0.0 – 0.3 |
|  | *Schistosoma mansoni* | 66 | 2.4 | 1.9 – 0.3 |
| Total |  | 183 | 6.6 | 5.7 – 7.6 |
| Poly-parasitism | Hookworm/*Entamoeba* spp. | 6 | 0.2 | 0.1 – 0.4 |
|  | Hookworm/*Schistosoma mansoni* | 3 | 0.1 | 0.0 – 0.3 |
|  | Hookworm/*Ascaris lumbricoides* | 3 | 0.1 | 0.0 – 0.3 |
| Total |  | 12 | 0.4 | 0.2 – 0.7 |

**Table 3.**
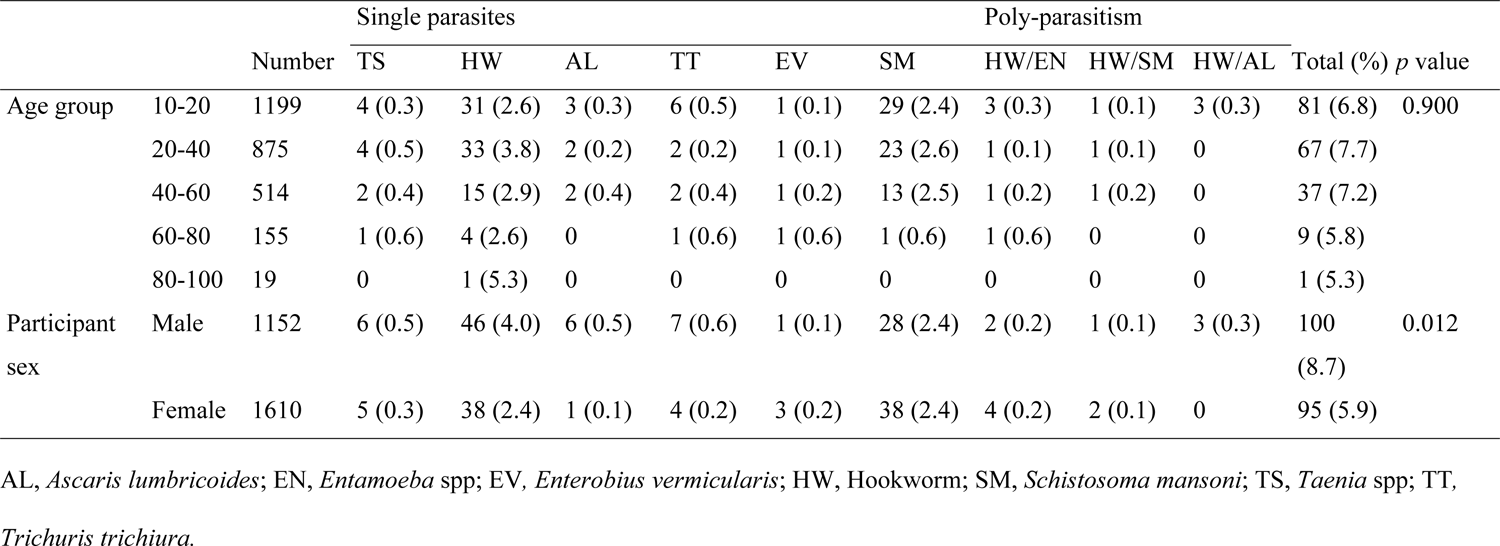
Prevalence of intestinal helminths by age and sex in the Chiparamba area of Chipata district, Eastern province, Zambia.

**Table 4.**
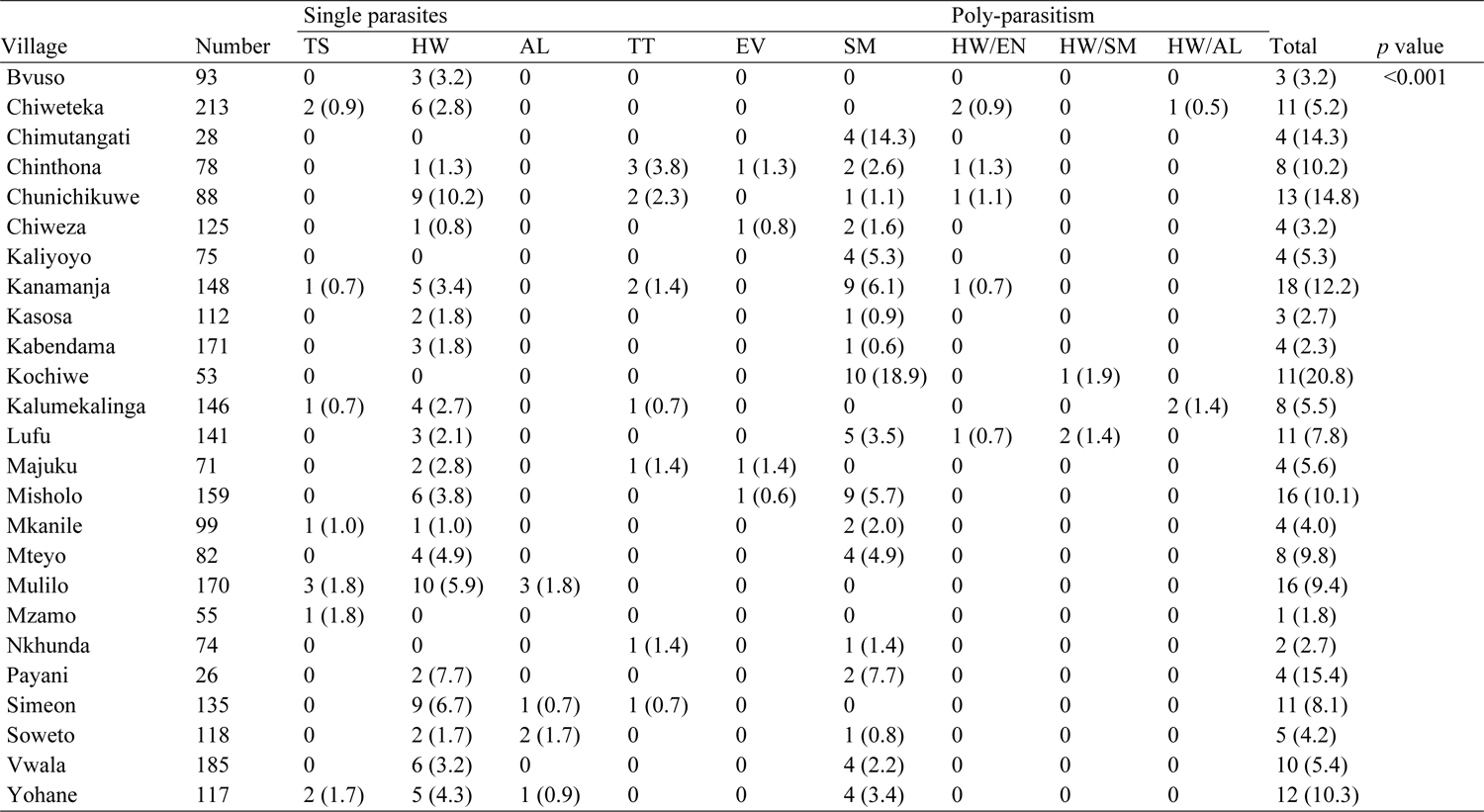
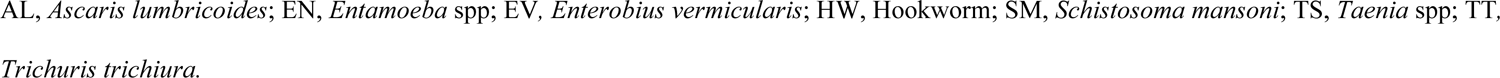
Prevalence of intestinal helminths by village in the Chiparamba area of Chipata district, Eastern province, Zambia.

| Village | Number | Single parasites |  |  |  |  |  | Poly-parasitism |  |  | Total | *p* value |
| --- | --- | --- | --- | --- | --- | --- | --- | --- | --- | --- | --- | --- |
|  |  | TS | HW | AL | TT | EV | SM | HW/EN | HW/SM | HW/AL |  |  |
| Bvuso | 93 | 0 | 3 (3.2) | 0 | 0 | 0 | 0 | 0 | 0 | 0 | 3 (3.2) | <0.001 |
| Chiweteka | 213 | 2 (0.9) | 6 (2.8) | 0 | 0 | 0 | 0 | 2 (0.9) | 0 | 1 (0.5) | 11 (5.2) |  |
| Chimutangati | 28 | 0 | 0 | 0 | 0 | 0 | 4 (14.3) | 0 | 0 | 0 | 4 (14.3) |  |
| Chinthona | 78 | 0 | 1 (1.3) | 0 | 3 (3.8) | 1 (1.3) | 2 (2.6) | 1 (1.3) | 0 | 0 | 8 (10.2) |  |
| Chunichikuwe | 88 | 0 | 9 (10.2) | 0 | 2 (2.3) | 0 | 1 (1.1) | 1 (1.1) | 0 | 0 | 13 (14.8) |  |
| Chiweza | 125 | 0 | 1 (0.8) | 0 | 0 | 1 (0.8) | 2 (1.6) | 0 | 0 | 0 | 4 (3.2) |  |
| Kaliyoyo | 75 | 0 | 0 | 0 | 0 | 0 | 4 (5.3) | 0 | 0 | 0 | 4 (5.3) |  |
| Kanamanja | 148 | 1 (0.7) | 5 (3.4) | 0 | 2 (1.4) | 0 | 9 (6.1) | 1 (0.7) | 0 | 0 | 18 (12.2) |  |
| Kasosa | 112 | 0 | 2 (1.8) | 0 | 0 | 0 | 1 (0.9) | 0 | 0 | 0 | 3 (2.7) |  |
| Kabendama | 171 | 0 | 3 (1.8) | 0 | 0 | 0 | 1 (0.6) | 0 | 0 | 0 | 4 (2.3) |  |
| Kochiwe | 53 | 0 | 0 | 0 | 0 | 0 | 10 (18.9) | 0 | 1 (1.9) | 0 | 11 (20.8) |  |
| Kalumekalinga | 146 | 1 (0.7) | 4 (2.7) | 0 | 1 (0.7) | 0 | 0 | 0 | 0 | 2 (1.4) | 8 (5.5) |  |
| Lufu | 141 | 0 | 3 (2.1) | 0 | 0 | 0 | 5 (3.5) | 1 (0.7) | 2 (1.4) | 0 | 11 (7.8) |  |
| Majuku | 71 | 0 | 2 (2.8) | 0 | 1 (1.4) | 1 (1.4) | 0 | 0 | 0 | 0 | 4 (5.6) |  |
| Misholo | 159 | 0 | 6 (3.8) | 0 | 0 | 1 (0.6) | 9 (5.7) | 0 | 0 | 0 | 16 (10.1) |  |
| Mkanile | 99 | 1 (1.0) | 1 (1.0) | 0 | 0 | 0 | 2 (2.0) | 0 | 0 | 0 | 4 (4.0) |  |
| Mteyo | 82 | 0 | 4 (4.9) | 0 | 0 | 0 | 4 (4.9) | 0 | 0 | 0 | 8 (9.8) |  |
| Mulilo | 170 | 3 (1.8) | 10 (5.9) | 3 (1.8) | 0 | 0 | 0 | 0 | 0 | 0 | 16 (9.4) |  |
| Mzamo | 55 | 1 (1.8) | 0 | 0 | 0 | 0 | 0 | 0 | 0 | 0 | 1 (1.8) |  |
| Nkhunda | 74 | 0 | 0 | 0 | 1 (1.4) | 0 | 1 (1.4) | 0 | 0 | 0 | 2 (2.7) |  |
| Payani | 26 | 0 | 2 (7.7) | 0 | 0 | 0 | 2 (7.7) | 0 | 0 | 0 | 4 (15.4) |  |
| Simeon | 135 | 0 | 9 (6.7) | 1 (0.7) | 1 (0.7) | 0 | 0 | 0 | 0 | 0 | 11 (8.1) |  |
| Soweto | 118 | 0 | 2 (1.7) | 2 (1.7) | 0 | 0 | 1 (0.8) | 0 | 0 | 0 | 5 (4.2) |  |
| Vwala | 185 | 0 | 6 (3.2) | 0 | 0 | 0 | 4 (2.2) | 0 | 0 | 0 | 10 (5.4) |  |
| Yohane | 117 | 2 (1.7) | 5 (4.3) | 1 (0.9) | 0 | 0 | 4 (3.4) | 0 | 0 | 0 | 12 (10.3) |  |

AL, *Ascaris lumbricoides*; EN, *Entamoeba* spp; EV, *Enterobius vermicularis*; HW, Hookworm; SM, *Schistosoma mansoni*; TS, *Taenia* spp; TT, *Trichuris trichiura*.

Regarding intestinal parasite infection per village, of the 25 villages sampled, 21 had participants with hookworm infection, while 18 villages had participants infected with *S. mansoni*. The number of villages with particular intestinal parasite infections is shown in (Fig 4). Entamoeba spp Schistosoma mansoni Enterobius vermicularis Trichuris trichiura Ascaris lumbricoides Hookworm spp. Taenia spp.

**Fig 4:**
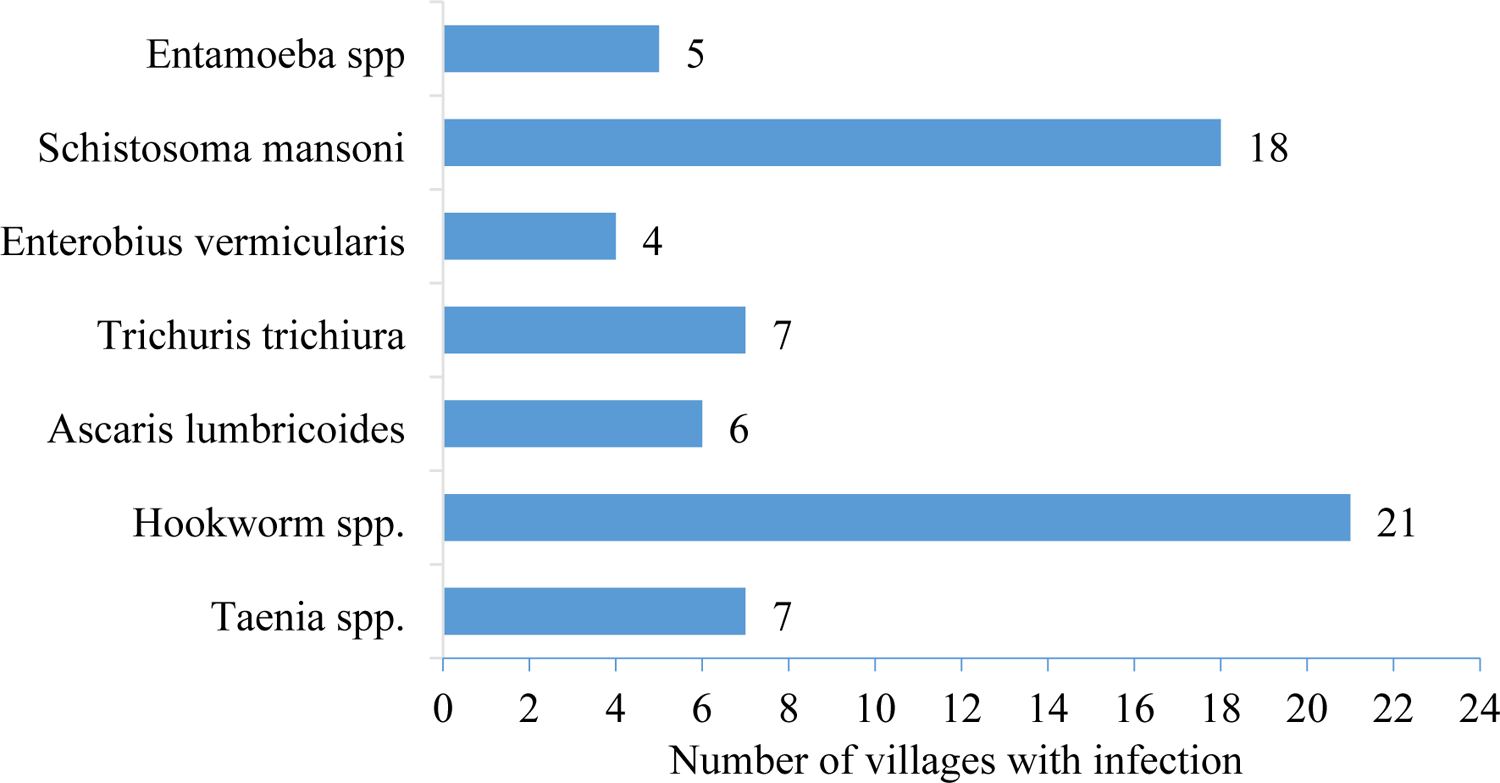
Intestinal parasites and number of villages with infection in the Chiparamba area of Chipata district, Eastern province, Zambia.

### Predictors of being positive for intestinal parasite infection

Using binary logistic regression, sex and village were significantly associated with intestinal parasite infection. The odds of male participants being positive for parasite infection were 1.5 times higher than those of females (95% CI 1.1 – 2.0, *p* = 0.008). The odds of being positive for intestinal parasite infection were significantly higher for Chiweteka, Chinthona, Kaliyoyo, Kabendama and Nkhunda villages than for Yohane village (Table 5).

**Table 5.** Predictors of a positive intestinal parasite result.

| Predictor variable |  | OR | 95% CI | *p value* |
| --- | --- | --- | --- | --- |
| Sex | Male | 1.5 | 1.1 – 2.0 | 0.008 |
|  | Female | Ref |  |  |
| Village | Bvuso | 1.4 | 0.4 – 5.1 | 0.658 |
|  | Chiweteka | 5.1 | 1.1 – 24.3 | 0.042 |
|  | Chimutangati | 3.4 | 0.9 – 13.3 | 0.080 |
|  | Chinthona | 5.4 | 1.5 – 19.7 | 0.011 |
|  | Chunichikuwe | 1.0 | 0.2 – 4.6 | 0.990 |
|  | Chiweza | 1.8 | 0.4 – 8.1 | 0.470 |
|  | Kaliyoyo | 4.0 | 1.1 – 14.1 | 0.030 |
|  | Kanamanja | 0.8 | 0.2 – 4.4 | 0.837 |
|  | Kasosa | 0.7 | 0.2 – 3.3 | 0.686 |
|  | Kabendama | 7.8 | 2.1 – 29.7 | 0.002 |
|  | Kochiwe | 1.5 | 0.4 – 6.0 | 0.552 |
|  | Kalumekalinga | 2.6 | 0.7 – 9.6 | 0.152 |
|  | Lufu | 2.0 | 0.4 – 9.0 | 0.392 |
|  | Majuku | 3.4 | 1.0 – 11.9 | 0.060 |
|  | Misholo | 1.0 | 0.2 – 4.9 | 0.961 |
|  | Mkanile | 3.3 | 0.9 – 13.0 | 0.083 |
|  | Mteyo | 2.5 | 0.7 – 9.2 | 0.153 |
|  | Mulilo | 0.0 | 0.0 | 0.997 |
|  | Mzamo | 0.8 | 0.1 – 5.1 | 0.841 |
|  | Nkhunda | 5.5 | 1.1 – 26.3 | 0.034 |
|  | Payani | 2.7 | 0.7 – 10.1 | 0.130 |
|  | Simeon | 1.4 | 0.3 – 6.0 | 0.660 |
|  | Soweto | 1.7 | 0.5 – 6.4 | 0.426 |
|  | Vwala | 2.9 | 0.8 – 10.9 | 0.113 |
|  | Yohane | Ref |  |  |

286 OR, odds ratio; Ref, reference standard.

### Intestinal helminthiasis due to taeniasis

Of the 2762 stool samples submitted, 241 (8.7%, 95% CI = 7.7 – 9.8) tested positive for taeniasis on copro Ag-ELISA, and 11 (0.4%) tested positive on microscopy. Overall, 241 (8.7%) participants tested positive for taeniasis on either copro Ag-ELISA or microscopy, with eleven (0.4%, 95% CI = 0.2 – 0.7) participants testing positive on both copro Ag-ELISA and microscopy examination. The majority (230, 95.4%, 95% CI = 92.0 – 97.4) were positive only by copro Ag-ELISA (Table 6).

**Table 6.** Prevalence of taeniasis based on copro Ag-ELISA and microscopic examination of stool samples in the Chiparamba area of Chipata district, Eastern province, Zambia.

|  | Number | Positive | Percent | 95%CI |
| --- | --- | --- | --- | --- |
| Copro Ag-ELISA | 2762 | 241 | 8.7 | 7.7 – 9.8 |
| Microscopy |  | 11 | 0.4 | 0.2 – 0.7 |
| *Results on parallel test with copro Ag-ELISA and microscopy* |  |  |  |  |
| Pos copro Ag-ELISA/Pos microscopy | 241 | 11 | 4.6 | 2.6 – 8.0 |
| Pos copro Ag-ELISA/Neg microscopy |  | 230 | 95.4 | 92.0 – 97.4 |

Ag, Antigen; CI, Confidence interval; ELISA, Enzyme-linked immunosorbent assay; Neg, Negative; Pos, Positive.

When we compared taeniasis prevalence by age group, it was observed that the 60-80 age group reported a higher prevalence by copro Ag-ELISA than those below 60 years and those above 80 years (Fig 5).

**Fig 5.**
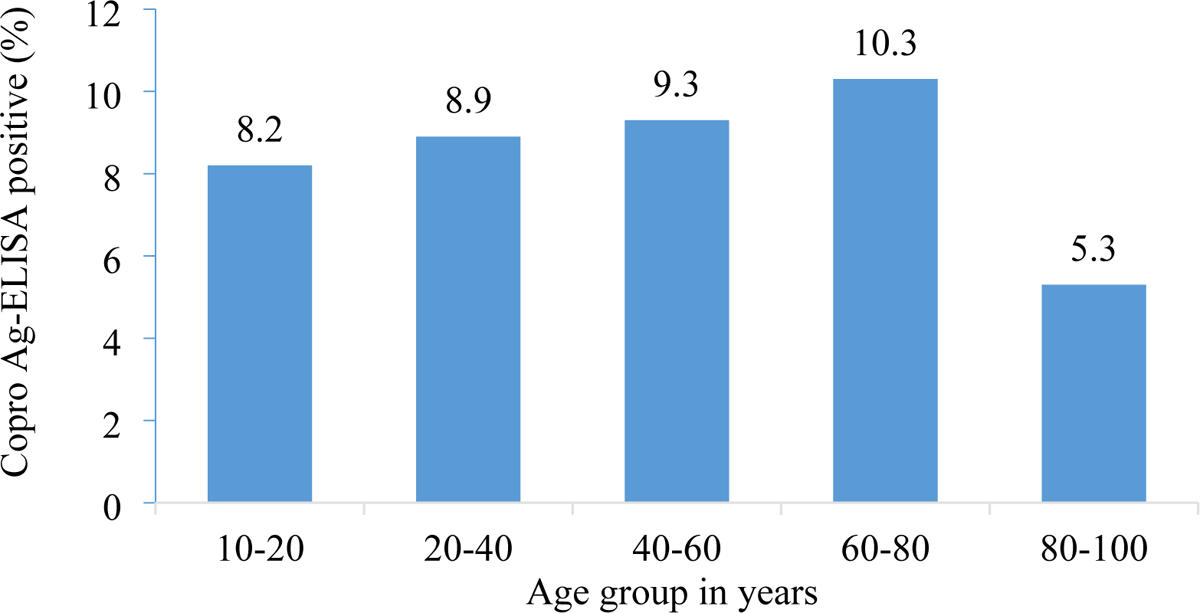
Taeniasis prevalence based on copro Ag-ELISA by age group in the Chiparamba area of Chipata district, Eastern province, Zambia.

No positive case on copro Ag-ELISA was reported from 6 villages, and based on microscopy examination of the stool, seven villages had at least one person with *Taenia* spp. eggs (Table 7). Tapeworm proglottids were obtained from one participant after treatment with 2 grams of niclosamide and purgation with magnesium sulfate, although these were not confirmed if they were for *T. solium*.

**Table 7.** Prevalence of taeniasis based on copro Ag-ELISA and microscopy by village in Chiparamba area of Chipata district, Eastern province, Zambia.

| Village | Number | Copro Ag-ELISA |  | *p* value | Microscopy |  | *p* value |
| --- | --- | --- | --- | --- | --- | --- | --- |
|  |  | Positive (%) | 95%CI |  | Positive (%) | 95%CI |  |
| Bvuso | 93 | 0 |  | <0.001 | 0 |  | 0.430 |
| Chiweteka | 213 | 35 (16.4) | 12.0 – 22.0 |  | 2 (0.9) | 0.2 – 3.3 |  |
| Chimutangati | 28 | 0 |  |  | 0 |  |  |
| Chinthona | 78 | 0 |  |  | 0 |  |  |
| Chunichikuwe | 88 | 0 |  |  | 0 |  |  |
| Chiweza | 125 | 3 (2.4) | 0.8 – 6.8 |  | 0 |  |  |
| Kaliyoyo | 75 | 2 (2.7) | 0.7 – 9.3 |  | 0 |  |  |
| Kanamanja | 148 | 25 (16.9) | 11.7 – 23.8 |  | 1 (0.7) | 0.13 – 3.8 |  |
| Kasosa | 112 | 13 (11.6) | 6.9 – 18.8 |  | 0 |  |  |
| Kabendama | 171 | 27 (15.8) | 11.1 – 22.0 |  | 0 |  |  |
| Kochiwe | 53 | 0 |  |  | 0 |  |  |
| Kalumekalinga | 146 | 14 (9.6) | 5.8 – 15.5 |  | 1 (0.7) | 0.1 – 3.8 |  |
| Lufu | 141 | 10 (7.1) | 3.9 – 12.6 |  | 0 |  |  |
| Majuku | 71 | 6 (8.5) | 4.0 – 17.3 |  | 0 |  |  |
| Misholo | 159 | 42 (26.4) | 20.1 – 33.8 |  | 0 |  |  |
| Mkanile | 99 | 8 (8.1) | 4.2 – 15.2 |  | 1 (1.0) | 0.2 – 5.5 |  |
| Mteyo | 82 | 8 (9.8) | 5.2 – 17.8 |  | 0 |  |  |
| Mulilo | 170 | 3 (1.8) | 0.6 – 5.1 |  | 3 (1.8) | 0.6 – 5.1 |  |
| Mzamo | 55 | 1 (1.8) | 0.3 – 9.6 |  | 1 (1.8) | 0.3 – 9.6 |  |
| Nkhunda | 74 | 10 (13.5) | 7.5 – 23.1 |  | 0 |  |  |
| Payani | 26 | 0 |  |  | 0 |  |  |
| Simeon | 135 | 2 (1.5) | 0.4 – 5.3 |  | 0 |  |  |
| Soweto | 118 | 6 (5.1) | 2.4 – 10.7 |  | 0 |  |  |
| Vwala | 185 | 21 (11.4) | 7.6 – 16.8 |  | 0 |  |  |
| Yohane | 117 | 5 (4.3) | 1.9 – 9.6 |  | 2 (1.7) | 0.5 – 6.0 |  |
| Total | 2762 | 241 (8.7) | 7.7 – 9.8 |  | 11 (0.4) | 0.2 – 0.7 |  |

Considering the results based on copro Ag-ELISA and microscopy examination, there was a statistically significant difference in the prevalence of taeniasis based on copro Ag-ELISA among the villages (*p* <0.001), with the highest prevalence reported in Misholo village at 26.4% (Fig 6, Table 7). However, sex and age were not significantly associated with the prevalence of taeniasis in our study area (Table 8).

**Fig 6.**
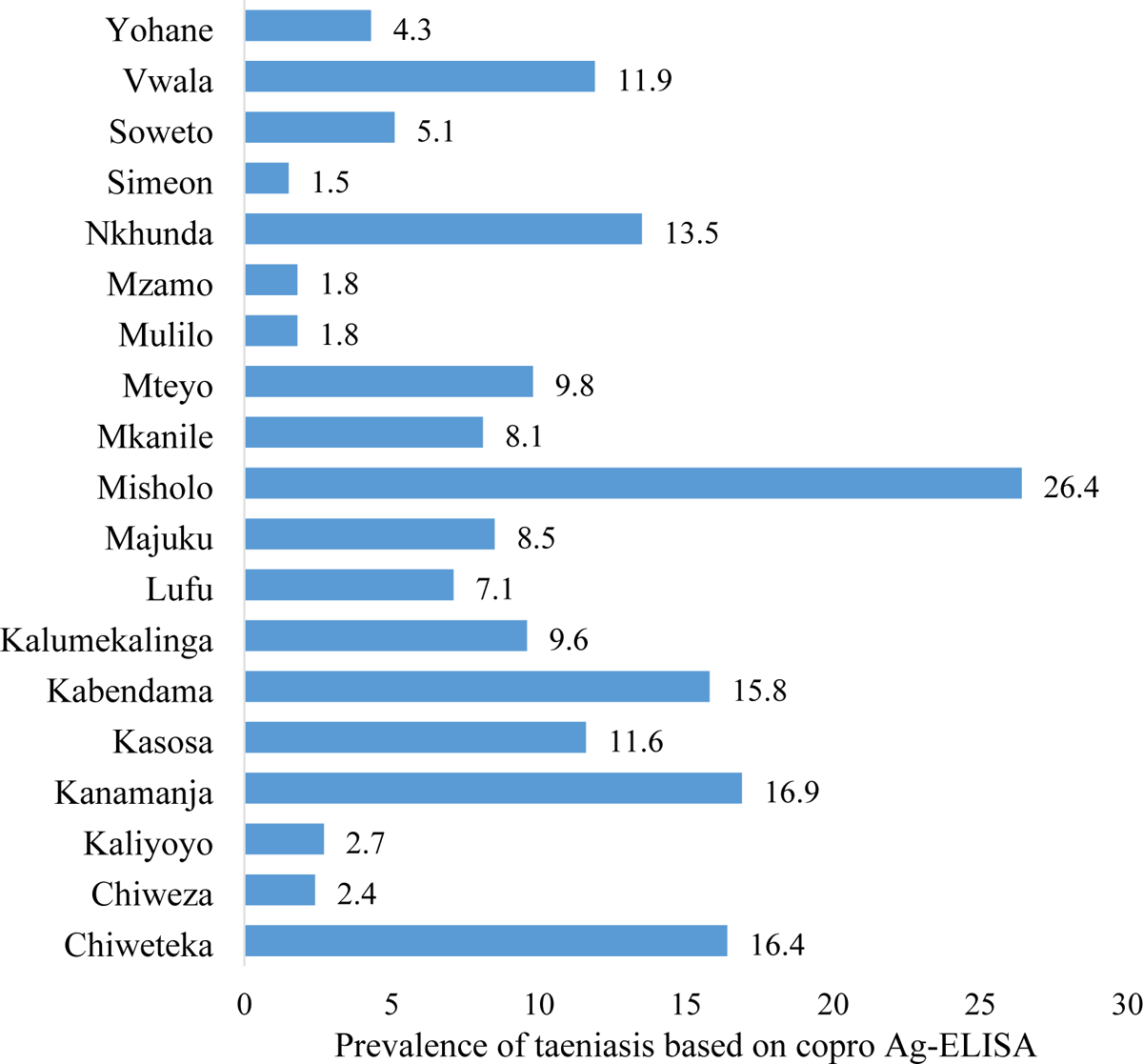
Prevalence of taeniasis based on copro Ag-ELISA by village in the Chiparamba area of Chipata district, Eastern province, Zambia.

**Table 8.** Comparison of taeniasis prevalence based on copro Ag-ELISA and microscopy with sex and age in the Chiparamba area of Chipata district, Eastern province, Zambia.

|  |  |  | Copro Ag-ELISA |  |  | Microscopy |  |  |
| --- | --- | --- | --- | --- | --- | --- | --- | --- |
|  |  | Number | Positive (%) | 95% CI | *p* value | Positive (%) | 95% CI | *p* value |
| Sex | Male | 1152 | 105 (9.1) | 7.6 – 10.9 | 0.539 | 6 (0.5) | 0.2 – 1.1 | 0.542 |
|  | Female | 1610 | 136 (8.4) | 7.1 – 9.9 |  | 5 (0.3) | 0.1 – 0.7 |  |
| Age group | 10-20 | 1199 | 98 (8.2) | 6.8 – 9.9 | 0.823 | 4 (0.3) | 0.1 – 0.8 | 0.764 |
|  | 20-40 | 875 | 78 (8.9) | 7.2 – 11.0 |  | 4 (0.5) | 0.2 – 1.2 |  |
|  | 40-60 | 514 | 48 (9.3) | 7.1 – 12.1 |  | 2 (0.4) | 0.1 – 1.4 |  |
|  | 60-80 | 155 | 16 (10.3) | 6.4 – 16.1 |  | 1 (0.6) | 0.1 – 3.5 |  |
|  | 80-100 | 19 | 1 (5.3) | 0.9 – 24.7 |  | 0 | 0.0 – 0.2 |  |

### Predictors of a positive taeniasis result

Using binary logistic regression, only village was found to be a significant predictor of being positive for taeniasis based on copro Ag-ELISA results, with nine villages having significantly higher odds compared to Yohane village (Table 9). No variables had significant odds of being positive on microscopy.

**Table 9.** Predictors of a positive taeniosis result on copro Ag-ELISA.

|  | Predictor variable | OR | 95% CI | *p* value. |
| --- | --- | --- | --- | --- |
| Village | Bvuso | 0.0 | 0.0 | 0.997 |
|  | Chiweteka | 6.9 | 2.1 – 23.1 | 0.002 |
|  | Chimutangati | 0.0 | 0.0 | 0.998 |
|  | Chinthona | 0.0 | 0.0 | 0.997 |
|  | Chunichikuwe | 0.0 | 0.0 | 0.997 |
|  | Chiweza | 0.9 | 0.2 – 4.6 | 0.918 |
|  | Kaliyoyo | 1.0 | 0.2 – 6.3 | 0.981 |
|  | Kanamanja | 7.3 | 2.1 – 24.9 | 0.002 |
|  | Kasosa | 4.9 | 1.4 – 17.7 | 0.015 |
|  | Kabendama | 7.0 | 2.1 – 23.7 | 0.002 |
|  | Kochiwe | 0.0 | 0.0 | 0.997 |
|  | Kalumekalinga | 3.7 | 1.0 – 13.2 | 0.046 |
|  | Lufu | 2.9 | 0.8 – 10.6 | 0.118 |
|  | Majuku | 3.4 | 0.8 – 14.2 | 0.088 |
|  | Misholo | 13.4 | 4.0 – 44.5 | <0.001 |
|  | Mkanile | 2.9 | 0.7 – 11.4 | 0.134 |
|  | Mteyo | 4.0 | 1.0 – 15.7 | 0.044 |
|  | Mulilo | 0.0 | 0.0 | 0.995 |
|  | Mzamo | 0.0 | 0.0 | 0.997 |
|  | Nkhunda | 5.8 | 1.5 – 22.0 | 0.009 |
|  | Payani | 0.0 | 0.0 | 0.998 |
|  | Simeon | 0.6 | 0.1 – 3.4 | 0.531 |
|  | Soweto | 2.0 | 0.5 – 8.2 | 0.335 |
|  | Vwala | 4.8 | 1.4 – 16.4 | 0.013 |
|  | Yohane | Ref |  |  |

337 OR, odds ratio; Ref, reference standard.

## Discussion

This study aimed to determine the prevalence of intestinal helminthiasis and taeniasis in a rural community known to have risk factors for *T. solium* in the eastern province of Zambia.

We found that intestinal helminth infections are endemic in our study area, with 7.1% of the participants infected with at least one intestinal helminth species. Among the nematode species, hookworm infection was the most predominant, followed by *A. lumbricoides* and *T. trichiura* infection. The prevalence of hookworms was similar to the 3.5% prevalence reported among the general population in the Southern province of Zambia [15]. However, the prevalence reported in our study was much lower than that reported in previous studies in Zambia. For instance, in the Luangwa, Kalabo, and Serenje districts of Zambia, hookworm prevalence ranging from 12% to 35% was reported [25]. Previous studies conducted in the central province of Zambia also reported a high hookworm infection prevalence ranging from 11% to 77% [26]. Hookworm infections have also been the predominant intestinal helminths reported in some studies conducted among the general population in Ethiopia [27], India [28] and South Africa [29]. However, in most studies among children, infection with *A. lumbricoides* has generally been higher than that with hookworms. A study conducted among children in Chililabombwe district and another conducted at a children’s hospital in Ndola on the Copperbelt province of Zambia showed *A. lumbricoides* infection to be more prevalent compared to hookworm and other nematodes [17,18]. Globally, it is also estimated that infection with *A*. *lumbricoides* has the widest distribution [30]. The overall prevalence of intestinal helminths in our study was lower than that reported in previous studies in Zambia, which reported numbers of up to 12 – 35% [15,17,25]. One reason could be that our sample size was much larger than that used in other studies and may therefore be more representative. The other reason is probably due to the annual mass drug administration (MDA), a deworming programme conducted by the Ministry of Health in conjunction with the Ministry of Education, leading to the assumption that these programmes have a positive effect on infection prevalence. Infection with helminths has been shown to affect the growth and development of children, and children with helminth infections are also more likely to be absent from school, hence affecting school attendance [6]. Subsequent continued annual treatment of school-aged children for soil-transmitted helminths has also contributed to a reduction in the number of eggs being expelled and the resulting prevalence [31,32].

As another parasitic worm infection, schistosomiasis is also endemic in Zambia [33]. The prevalence of *S. mansoni* within our study was similar to the overall prevalence of 2.9% found in the Southern province of Zambia [15], which can be classified as low prevalence according to WHO grading [6]. However, a higher prevalence of up to 73% for *S. mansoni* has been reported in the Western province [34]. In our study, schistosomiasis was more prevalent in villages that were located near streams. This observation may indicate a lack of sufficient sanitary facilities and the use of contaminated water for domestic and recreational activities [15,35,36]. *S. mansoni* has also been associated with long-term health problems. For example, in the northwestern province of Zambia, cases of periportal liver fibrosis among rural children were reported [37], and in the western province, cases of periportal liver fibrosis and main portal branch fibrosis were reported among children [35]. This calls for disease control managers and policy makers in the country to devise new control strategies and regular disease surveillance programmes through multisectoral collaborations, which contribute towards having successful MDA programmes informed by disease distribution patterns.

With effective knowledge sharing, improved sanitary conditions and deworming, control of intestinal helminth infections can be achieved. Deworming, which is safe, effective, and inexpensive, has been shown to improve the growth and development of children [3]. These MDA programmes need to be sustained and should focus more on high prevalence villages, including targeting adults [15].

Hyperendemicity for *T. solium* taeniasis in an area is defined as a point prevalence rate greater than 1% [38]. Our study community, with an estimated prevalence of 8.7% by copro Ag-ELISA, was thus hyperendemic to taeniasis. The infection displayed great heterogeneity between villages, as seen from the significant variability in proportions between villages ranging from 0 – 26%. Similar variations were also reported in studies conducted in Peru and Guatemala using copro Ag-ELISA [39,40]. The variation between villages could be due to differences in the consumption of infected pork, cooking habits or household dietary habits. Further studies are required to understand the causes of this variation.

The prevalence of taeniasis by copro Ag-ELISA in our study area was much higher than that found using stool microscopy. Copro Ag-ELISA has a sensitivity that is 2 – 10 times higher than that of microscopy, which could explain the difference in the two prevalences [41]. However, we are cautious in interpreting the high proportion on copro Ag-ELISA because the test cannot differentiate between *T. solium* and *Taenia saginata* infections, as it is only genus specific [41]. It is therefore possible that the proportions of people being positive for *T. solium* could have been overestimated. Nonetheless, based on samples from within the Eastern province of Zambia, Bayesian modelling estimating the test characteristics of coprology, copro Ag-ELISA and PCR for the diagnosis of taeniasis yielded estimates for sensitivity and specificity of 84.5% and 92% for copro Ag-ELISA [42]. Despite cattle farming being highly practiced and the consumption of beef being high, bovine cysticercosis in Zambia has thus far only been reported in the central and southern provinces [43]; hence, the risk for coinfection with *T. saginata* in our study area was low.

The overall taeniasis positivity determined by copro Ag-ELISA in this study is higher than the prevalence of 6.3% reported in Petauke district within the Eastern province of Zambia but lower than the 11.9% reported in the neighboring district of Katete [7,14]. It was also higher than the 5.2% reported in Tanzania [44] and 1.4% in Rwanda [45] but lower than the 19.7% reported in Kenya [46] and 23.4% in the Democratic Republic of Congo [47]. As a potential risk factor for *T. solium* taeniasis, consumption of poorly cooked pork and pork with cysts was reported to be practiced in our study area, with men in particular being the most exposed to undercooked meat within the Eastern province of Zambia, translating into a higher taeniasis prevalence among them [14]. Nevertheless, considering the predictors of taeniasis, we did not find any association between positivity for taeniasis and age or sex in our study area. This was also the case in comparable studies conducted within the Eastern province of Zambia [14], in Tanzania [44] and in studies conducted in other countries in Latin America [38–40,48,49]. In contrast, in a study conducted in Congo, age was found to be significantly associated with taeniasis, and the highest positivity was found in subjects belonging to the 5 – 10 years age group [47]. This association was also observed in studies conducted in Peru and Guatemala [39,40]. Our study excluded participants aged younger than 10 years, so we cannot predict what the outcome would have been had these been included. Nonetheless, it was observed that taeniasis prevalence both on copro Ag-ELISA and microscopy was lower in those aged below 40 years. However, since all age groups are affected by taeniasis, intervention strategies such as MDA should target the entire population [5].

In addition to resulting in intestinal taeniasis and preserving the life cycle of *T. solium*, a tapeworm carrier also poses a risk for the development of human cysticercosis, resulting in even more severe health conditions, such as epilepsy. Transmission occurs either through autoinfection or transmission to other community members by ingesting infective eggs/proglottids. *T. solium* tapeworm proglottids were, for example, recovered from one of the taeniasis-positive participants whose trade was mainly herding cattle in the fields. With the non-availability of latrines in the fields and, in general, the non-use of latrines within the province, open defaecation is practiced [50,51]. Environmental contamination with tapeworm eggs is thus inevitable, leading to an increased exposure risk to infective eggs for both humans and pigs. The latter could also explain the high taeniasis prevalence recorded in our study community [14].

While the major strength of our study was the large sample size used to estimate the prevalence of intestinal helminths, including taeniasis, compared to previous studies within the Eastern province of Zambia, the study had some limitations. Among them were the few variables linked to people, pigs and the environment obtained to ascertain risk factors for intestinal helminthiasis and taeniasis. Another limitation is that quantification of the helminth eggs was not performed, and therefore, the intensity of helminth infection in our study population could not be ascertained. Additionally, participants were not asked if they had taken any anti-helminthic medication in the past three months prior to our study to have a true estimate of intestinal helminthiasis prevalence. The fact that only one stool sample was collected instead of three consecutive samples for microscopy could have led to the underestimation of intestinal parasite infections reported in our study.

## Conclusion

Intestinal helminth infections, including taeniasis, most likely *T. solium* taeniasis and *S. mansoni*, are hyperendemic in the Chiparamba area of the Chipata district in the Eastern province of Zambia. While the estimates of prevalence may be lower in our study area compared to other districts within Zambia, they are still high enough to warrant public health interventions for surveillance and control. More work has to be done to ensure the enforcement of public health interventions for the control and elimination of these infections in rural parts of Zambia.

## Data Availability

The data supporting the conclusions of this article are included within the article. Raw datasets used and/or analysed during the current study are available from the corresponding author on reasonable request.

## Acknowledgements

We would like to acknowledge the study participants from the Chiparamba community and the nurses, clinicians and community health workers from the Chiparamba clinic for making this study possible. We would also like to acknowledge Gasiano Hapooma and Catherine Hatuma, the Laboratory Technicians from the Provincial Livestock and Fisheries Office, Chipata, for helping with sample processing during field work.

## Supplement

**S1 Fig.**
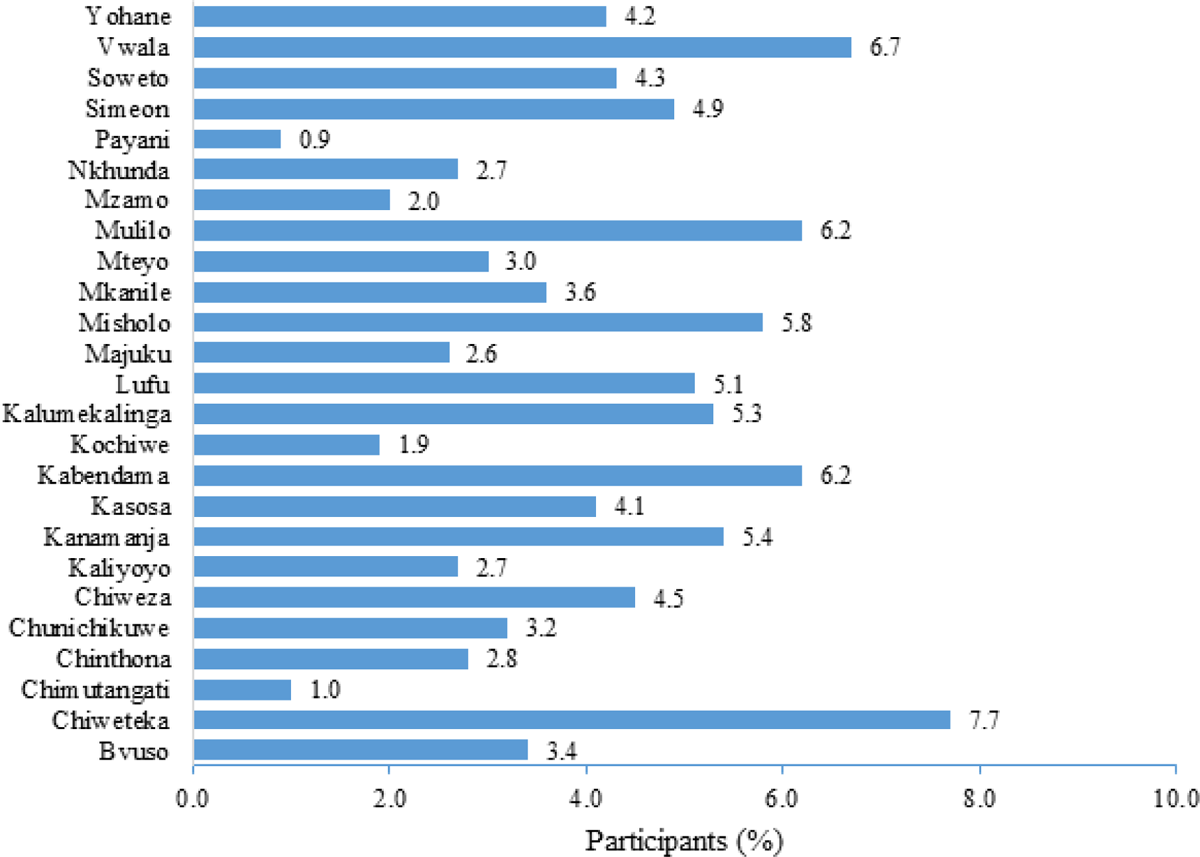
Percent distribution of participants by village in the study area. (TIF)

**S1 Table.** Strobe checklist. (DOCX)

## Author contributions

**Conceptualization:** Gideon Zulu, Chummy Sikasunge, Kabemba E. Mwape, Andrea S. Winkler, Isaac K. Phiri.

**Data curation:** Gideon Zulu.

**Formal analysis:** Gideon Zulu, Martin Simunza.

**Funding acquisition**: Chummy Sikasunge, Andrea S. Winkler.

**Investigation:** Gideon Zulu, Alex Hachangu, Maxwell Masuku, Mwelwa Chembensofu.

**Methodology:** Gideon Zulu, Chummy Sikasunge, Tamara M. Welte, Kabemba E. Mwape, Andrea S. Winkler, Isaac K. Phiri.

**Project administration:** Gideon Zulu, Chummy Sikasunge.

**Resources**: Chummy Sikasunge.

**Software**: Gideon Zulu, Martin Simunza,

**Supervision**: Andrea S. Winkler, Isaac K. Phiri.

**Validation**: Gideon Zulu, Tamara M. Welte, Kabemba E. Mwape, Andrea S. Winkler, Isaac K. Phiri,

**Visualization**: Gideon Zulu, Isaac K. Phiri.

**Writing** – original draft: Gideon Zulu

**Writing – review & editing**: Gideon Zulu, Chummy Sikasunge, Tamara M. Welte, Martin Simunza, Dominik Stelzle, Veronika Schmidt, Alex Hachangu, Wilbroad Mutale, Maxwell Masuku, Mwelwa Chembensofu, Clarissa Prazeres da Costa, Kabemba E. Mwape, Andrea S. Winkler, Isaac K. Phiri.

